# Tracking SARS-CoV-2 genomic variants in wastewater sequencing data with *LolliPop*

**DOI:** 10.1101/2022.11.02.22281825

**Authors:** David Dreifuss, Ivan Topolsky, Pelin Icer Baykal, Niko Beerenwinkel

**Affiliations:** Department of Biosystems Science and Engineering, ETH Zurich, CH-4058 Basel, Switzerland; SIB Swiss Institute of Bioinformatics, CH-1015 Lausanne, Switzerland

## Abstract

During the COVID-19 pandemic, wastewater-based epidemiology has progressively taken a central role as a pathogen surveillance tool. Tracking viral loads and variant outbreaks in sewage offers advantages over clinical surveillance methods by providing unbiased estimates and enabling early detection. However, wastewater-based epidemiology poses new computational research questions that need to be solved in order for this approach to be implemented broadly and successfully. Here, we address the variant deconvolution problem, where we aim to estimate the relative abundances of genomic variants from next-generation sequencing data of a mixed wastewater sample. We introduce *LolliPop*, a computational method to solve the variant deconvolution problem. *LolliPop* is tailored to wastewater time series sequencing data and applies temporal regularization in the form of a fused ridge penalty. We show that this regularization is equivalent to kernel smoothing and that it makes abundance estimates robust to very high levels of missing data, which is common for wastewater sequencing. We use the bootstrap to produce confidence intervals, and develop analytical standard errors that can produce similar confidence intervals at a fraction of the computational cost. We demonstrate the application of our method to data from the Swiss wastewater surveillance efforts as well as on simulated data.

**Author Summary:** Wastewater-based epidemiology has become a valuable tool for tracking viruses like SARS-CoV-2 across entire communities. Sequencing wastewater can reveal which viral variants are circulating, offering early and unbiased insights into variant dynamics. A central challenge is to infer the relative abundances of these variants from observed mutation data. This task is complicated by the fact that variant profiles can be highly similar, and the data is often noisy with many missing values, especially when the incidence of the pathogen is low. We developed *LolliPop*, a statistical method that leverages the time series structure of wastewater data to robustly deconvolve variant abundances and compute fast confidence intervals. Using both simulated data and real data from the Swiss national variant monitoring, we show that *LolliPop* is accurate and robust to high levels of missing data.

## Introduction

During the COVID-19 pandemic, genomic surveillance has been applied at an unprecedented scale to support various national efforts in containing outbreaks [1]. In this context, wastewater monitoring has seen its broadest and most successful application: PCR-based surveillance of total viral load has been deployed successfully in a large number of surveillance campaigns, some of which are now complementing this piece of information with next-generation sequencing (NGS) data to distinguish different genomic variants of SARS-CoV-2 [2]. As genomic analysis is extended from clinical samples to samples from wastewater treatment plants (WWTPs), new statistical and computational research questions arise: the samples are mixtures of heterogeneous, possibly highly diverse RNA molecules, for which the traditional approach of reconstructing consensus sequences is not suitable [1]. Beyond SARS-CoV-2, wastewater-based epidemiology (WBE) is becoming a central tool for pathogen surveillance in general [2]. WBE has been shown to inform on the infection dynamics of pathogens for which clinical testing is usually low, such as influenza and respiratory syncytial virus [3,4]. It is therefore pressing that the computational challenges of analyzing mixed, heterogeneous wastewater sequencing data be addressed.

Most of the existing viral genomic data analysis pipelines and tools were designed for clinical samples and rely on classifying the majority variant of each sample from the consensus sequence of the read alignment [5–7]. One of the main challenges in analyzing wastewater-derived NGS data is the loss of information about the full viral haplotype, i.e. which mutations occur together on the same RNA molecule. This loss can result from fragmentation of the genetic material and from sequencing protocols, which often rely on tiling amplicon PCR. In addition, the sequencing data exhibits very high levels of noise, because from a large pool of raw wastewater, extreme downsampling steps (grab or composite sampling, filtering, random reverse transcription, etc.) are followed by extreme amplification steps (PCR). This in turn can result in high levels of missing data. Some tools have been developed to increase sensitivity in the detection of variants, for example, by searching for co-occurring mutations on the same read, which has been shown to improve early detection of variants [8].

Beyond early detection, quantitative estimation of the relative abundances of different variants from wastewater is a very important endeavor. The growth rate of a new variant can inform on its fitness advantage relative to the dominating strain [9], and hence on its predicted impact on the infection dynamics. The fitness advantage is an epidemiologically important parameter that can be estimated accurately from wastewater samples using either specific PCR-based assays [10,11] or NGS data [8], while using far fewer samples as compared to using clinical data, provided that accurate quantification of the variant relative abundances can be made. For practical planning and policy making, it is therefore crucial to have accurate and time-efficient methods for estimation of the relative abundances through time of variants from wastewater NGS data, including reliable measures of uncertainty.

Prior research has shown that the relative abundance of an emerging variant can be quantified by averaging over a set of mutations that is unique for this variant [8]. However, as the number of variants grows, shared mutations cannot generally be discarded as finding sets of unique, characteristic mutations for each variant quickly becomes impossible. To address this limitation, some methods that take into account the correlation structure of mutations between different variants have been developed [12–15]. However, with these methods, quantification of uncertainty is done on the basis of the computationally intensive bootstrap, which can be prohibitive when faced with large amounts of data or limited computational resources (as is often the case in real-world surveillance settings). It is also not guaranteed in general that these methods fare well with high levels of missing data, which are common in environmental sampling due to low concentrations and high PCR inhibition [16].

Here, we introduce a new method for estimating variant relative abundances from wastewater sequencing data, named *LolliPop*. Tailored to time-series data, our method estimates the time course of relative abundances of all variants in the mixed sample. We employ temporal regularization to make this approach robust to high levels of missing data, which are frequent in wastewater sequencing. As an alternative to confidence bands based on the bootstrap, we derive analytical methods to compute asymptotic confidence bands, which provides a 30-fold speedup. We evaluate our method on wastewater data with high rates of missing values and compare the output to estimates derived from matched clinical data [8], as well as on simulated data. *LolliPop* is currently used for the Swiss wastewater monitoring program^1^.

## Methods

We model observed mutation frequencies in wastewater samples as a linear combination of variant profiles. To address challenges such as high noise, missing data, and similarity between variant profiles, we apply temporal regularization in the form of a fused ridge penalty. This leads to a regularized loss minimization framework for deconvolving relative variant abundances over time. We provide both bootstrap-based and analytical confidence intervals, incorporating adjustments for overdispersion and using logit reparametrization to ensure valid bounds. We evaluate the method on real wastewater sequencing data from the Swiss national surveillance program and on simulated datasets designed to assess robustness across varying levels of missing data and variant similarity.

### Variant deconvolution

We consider an ordered collection of samples, taken at (not necessarily evenly spaced) timepoints *t* = 1, …, *T*. Each studied variant *v* ∈ {1,…, *V*} carries a subset of the mutations *m ∈* {1,…, *M*} relative to a fixed reference strain. Let *X* ∈ {0, 1} ^*M× V*^ be the design matrix of variant definitions, i.e., *X*_*m,v*_ = 1 if variant *v* bears mutation *m*, and *X*_*m,v*_ = 0 otherwise. Let *y*_*t*_ *∈* [0, 1]^*M*^ be the observed mutation frequency vector at time *t*, where the entries are the observed proportions of reads from the wastewater sequencing experiment supporting a certain mutation *m*. We are interested in *b*_*t*_ ∈ [0, 1]^*V*^, ||*b*_*t*_ ||_1_ = 1, the relative variant abundances of all variants at each time point *t*. We make the assumption of a linear probability model, such that at time *t*, the expected proportion of reads with a given mutation is a linear combination of the relative variant abundances:

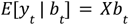

#### Definition (Variant Deconvolution Problem)

For given variant definitions *X* and a time series of mutation frequencies *y*_1_, …, *y*_*T*_, the variant deconvolution problem is to find the relative variant abundances *b*_1_, …, *b*_*T*_ in the population of the catchment area of the WWTP, such that *y*_*t*_ = *Xb*_*t*_ for all time points *t*.

As finding the exact relative variant abundances in the population is not possible due to the randomness of the data-generating process and model misspecification, we relax the problem to finding a best approximation. Solving the variant deconvolution problem is then performed by choosing a loss function *L* and optimizing:

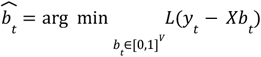

In the following, we use the soft *l* loss (SL1)

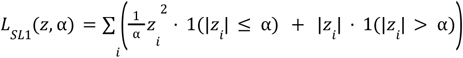

which interpolates between the least squares (LS) loss 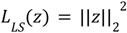 and the least absolute deviation loss *L*_*LAD*_ (*z*) = ||*z*||_1_ and is a common choice in robust statistics. Here, *α* is a hyperparameter controlling the tradeoff between robustness and efficiency.

### Temporal regularization

Wastewater sequencing data typically displays high noise levels, possibly leading to dropouts (i.e. missing values, genomic regions with no coverage). In the worst (and common) case entire rows corresponding to missing values of *y*_*t*_ are excluded and the variant design matrix *X* is rank deficient. In such a case, the variants relative abundances *b*_*t*_ are not identifiable without further assumptions. A common regularizer in regression settings where the design matrix is singular (or has a prohibitively high condition number) is the so-called ridge penalty[17]. We do not apply a ridge penalty to the magnitude of the relative abundance vectors, but we further assume temporal continuity of the variant abundances and introduce a fused ridge penalty on the difference between relative abundance vectors of different timesteps. We formulate the quadratic penalty 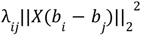 where *λ*_*ij*_ controls the penalization for different time differences between *b*_*i*_ and *b*_*j*_, and the penalization is distributed on the entries of *b*_*i*_ − *b*_*j*_ so as to avoid variants with a high count of defining mutations being less penalized. We extend the least square loss (SL1 loss with *α* = 1) with the ridge loss as follows,

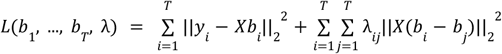

To minimize this loss function, we compute the gradient w.r.t *b* and set it to zero such that

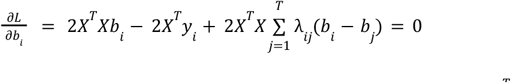

Where we assumed, without loss of generality, that *λ*_*ij*_ = *λ*_*ji*_. We define 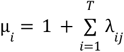 to obtain

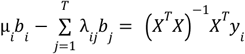

We define the doubly stochastic symmetric matrix

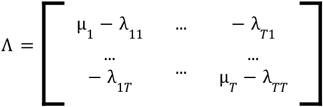

If *Λ* is full rank, which is easy to verify for example if *λ*_*ij*_ *> λ*_*ik*_ *∀* |*k − i*| *>* |*k − j*| (meaning that the penalization should encode a smoothness constraint), we obtain the solution

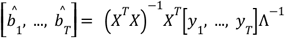

With the common convention of nonnegative penalties *λ*_*ij*_, we have that *Λ*^−1^ is also doubly stochastic. We remark that the solution is equivalent to solving the square loss solution with additional simultaneous kernel smoothing using the kernel function *k*(*i, j*) = [*Λ*^*−*1^]_*ij*_. Instead of specifying the *λ*_*ij*_ penalties, we therefore rather specify the more interpretable kernel function *k*(*i, j*), a non-negative, non-increasing function of *i − j*. Extending equation 1 we have:

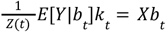

where *Y ∈* [0, 1]^*M×T*^ = [*y*_1_, …, *y*_*T*_], *k*_*t*_ = [*k*(*t, t*_1_),…, *k*(*t, t*_*T*_)]^*⊤*^ and 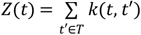. In *Lollipop*, we use the the Gaussian kernel 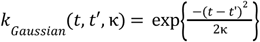 with bandwidth hyperparameter κ, which is a common choice in nonparametric statistics.

### Solving the deconvolution problem

With observed mutation frequencies *Y* and known variant definitions *X* as input, we solve the deconvolution problem for a given kernel *k*(*t, t*′) and loss function *L*(*z*) by finding 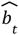 as

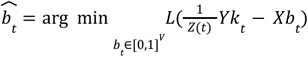

To numerically solve this optimization problem, we use routines from the Python scientific computing library *Scipy [18]*. In general we use the Trust Region Reflective method [19], but for *α* = 1 we can switch to using the LS loss function along with Scipy’s faster non-negative Least Squares solver [20].

### Confidence intervals

To use WBE for robust decision making, it is essential to provide estimates of the uncertainty in the prediction of relative variant abundances. We pursue two different strategies for computing confidence intervals: one based on the bootstrap and one based on an analytical approximation to the standard errors.

### Bootstrapping

A popular strategy to compute confidence bands is to use the non-parametric bootstrap [21]. Here, we construct *B* bootstrap samples of the whole time series, by resampling *M* mutation indices from *m ∈* 1, …, *M* with replacement. Each bootstrap sample is then processed by deconvolution and smoothing, resulting in *B* time series of dimensionality *V*. For each relative variant abundance *v ∈* 1, …, *V*, confidence intervals are constructed at each timepoint *t* from the empirical quantiles of the bootstrap samples.

### Asymptotic confidence intervals

We assume that at a given time *t*, the proportion *y*_*t,m*_ of reads supporting mutation *m* follows a *Binomial*/*n* distribution with parameter π_*m*_, i.e.,

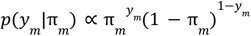

Using the linear probability model (Eq. 1), we have π_*m*_ = [*Xb*]_*m*_. We additionally assume conditional independence of the mutation proportions such that 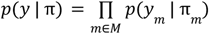. We thus obtain the log-likelihood:

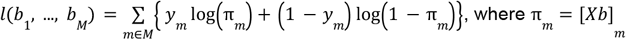

Differentiating twice the log-likelihood summands, we find the Fisher information matrix (see Supplementary 1)

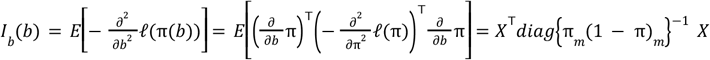

We extract the asymptotic standard errors:

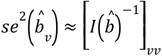

which are then used to construct Wald confidence intervals [22]. Here, a pseudofraction is added to the entries of *b* to avoid division by zero when computing the asymptotic standard errors.

### Logit reparametrization

To ensure that the confidence bands stay confined to the [0,1] interval, we compute the asymptotic standard errors and Wald confidence intervals on the logit scale 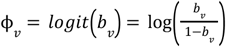, before projecting them back to the linear scale. We compute the inverse of the Fisher information matrix of ϕ by using the Delta method,

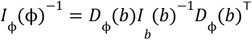

where *D*_*ϕ*_ (*b*) is the Jacobian of the transformation, such that

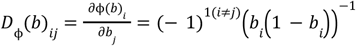

Here again, a pseudofraction is added to the entries of *b* to avoid division by zero.

### Overdispersion

The likelihood model we use for deriving asymptotic confidence intervals might insufficiently capture the noise of the observed data. We account for such overdispersion (and underdispersion) by following a quasilikelihood approach [23]. We compute the ratio of observed versus expected average square deviations of the observed data from the fitted values. This ratio is then taken as an overdispersion factor. At a given time *t*, the Wald confidence intervals are adjusted by adjusting the asymptotic standard errors for *b*_*t*_ :

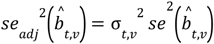

We build on the moment-based estimator of the dispersion factor for generalized linear models [23]

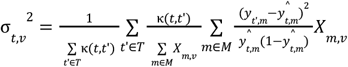

### Implementation and availability

The methods we present here are implemented in the Python package *LolliPop*, which takes as input a tabular file of observed mutation frequencies and variant definitions, performs simultaneous kernel smoothing and deconvolution using numerical optimization, and produces confidence intervals. *LolliPop* is available on Github (https://github.com/cbg-ethz/lollipop) and as a Bioconda package. All data and code used to produce the results presented in this article is available at https://doi.org/10.5281/zenodo.15277339.

### Processing of wastewater sequencing data

We used the wastewater sequencing data from the Swiss surveillance project reported in [8]. The dataset contains 1295 NGS datasets from longitudinal samples collected at six major WWTPs in Switzerland, sampled daily between January 2021 and September 2021. We defined the variants of concern (VOCs) B.1.1.7 (Alpha), B.1.351 (Beta), P.1 (Gamma), B.1.617.1 (Kappa), and B.1.617.2 (Delta) by querying the mutations present in ≥80% of the clinical sequences defining these variants on Cov-Spectrum [24] and supported by at least 100 clinical sequences. We then called these mutations in the wastewater samples from pileups of the read alignments. We deconvolved using different hyperparameter values (see below). We computed Wald confidence intervals adjusted for overdispersion with logit reparametrization, as well as bootstrap-based confidence intervals (1000 bootstrap samples).

### Comparison to clinical data

Using the LAPIS API of Cov-Spectrum [24], we retrieved counts of sequenced SARS-CoV-2 PCR-positive clinical samples for Switzerland, stratified by submitting lab, canton, and inferred variant. We restricted the data to samples from the large clinical testing company Viollier, where the PCR-positive samples are randomly subsampled before being sent for sequencing. We compare each WWTP to the clinical data from the canton it is located in. For the Berne WWTP of Laupen, we compare to an aggregate of the clinical sequences from both the cantons of Bern and Fribourg, as the catchment area is split between those two cantons [8].

### Simulations

As a matrix inverse problem, the variant deconvolution problem can be sensitive to collinearity in the variant definition matrix, which is determined by the genetic similarity between variants and exacerbated by noise and missing values. To assess the robustness of *Lollipop* to the degree of similarity between variants and the fraction of missing value in the data, we simulated wastewater sequencing data for a range of different scenarios. For timesteps *t ∈* 1,…, *T* and variants *v ∈* 1,…, *V*, we generated deterministic time series of variant relative abundances from a multinomial logistic growth model

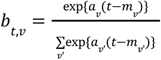

where the mixing of variants is controlled by parameter vectors *a* and *m*, which control the fitness and introduction times of the different variants. The expected frequencies of mutations at time *t* are computed as

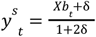

where *X* is the variant definition matrix and *δ* denotes the per-base error probability in the sequencing process. The read depth at position *m* and time *t* is then sampled as

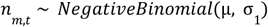

where μ and *σ*_1_ are the expected value and overdispersion parameter, respectively, of the read depth in covered regions. To simulate levels of dropouts higher than expected from this model we further randomly set entries *n*_*m,t*_ to zero with probability *p*. The observed mutation frequency at position *m* and time *t* is then sampled as

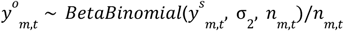

with expected value 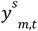 and overdispersion parameter *σ*_2_.

### Simulation of Delta taking over Alpha

To assess the efficacy of our method to track the important situation where a new variant displaces the currently circulating one over time, we generated a 60-timestep time series of Delta taking over Alpha at the logistic growth rate of 0.1/day. We used the variant definitions generated from Cov-Spectrum [24] using the toolkit from COJAC [8]. We set μ = 1000, *σ*_1_ = 10, *δ* = 0. 0025 and *σ*_2_ = 1000. To assess the robustness of our method to varying levels of noise, we varied the dropout probability *p* using 10 values linearly spaced between 0 and 0.99. These simulated datasets were deconvolved using the SL1 (*α* = 0. 135) loss, as well as with the LS (*α* = 1) loss to assess the robustness added by the SL1 loss. We varied the bandwidth κ of the kernel using 10 values linearly spaced between 0 and 60. We compared the deconvolved value to the known groundtruth and computed their squared correlation coefficient R^2^.

### Simulation of a mixture of Omicron subvariants

To further test our method in a more complicated situation where multiple related variants co-circulate, we generated another 60-timestep time series of a mixture of highly similar Omicron subvariants BA.2, BA.5, BA.2.75, BQ.1.1 and XBB. We set the other parameters of the simulation to the same values. We deconvolved using the same parameters and compared the results to the ground truth similarly.

### Simulation of a mixture of artificial closely related variants

Finally, we generated an artificially adversarial 60-timestep time series of a mixture of 5 highly related variants. Their definitions were generated by taking all size 4 subsets of a set of 5 mutations. This ensures no mutation is exclusive to any single variant, but that each of them is found in 4 of the 5 variants. Thus, each pair of variants shares all but one mutation. We set the rest of the parameters to the same values as before, deconvolved using the same procedure, and assessed the results similarly.

### Hyperparameters

We assessed the sensitivity of our deconvolution method to hyperparameter choice both on simulated and on real wastewater data. For the simulated data, we assessed the root mean square error of the deconvolution of a simulated mixture of 5 Omicron subvariants accross a grid of values for the smoothing bandwidth κ parameter and the *α* parameter (controlling the breakpoint between *l*_2_ and *l*_1_ loss) in (κ, *α*) *∈* [0, 30] *×* [0. 01, 1. 0], for different levels of missing data.

For the real wastewater sequencing data, we analyzed the two biggest WWTPs (Zurich and Vaud) using a similar grid of hyperparameters. For each deconvolved dataset, we linearly regressed the relative abundances of the different variants in clinical sequencing on the relative abundances in wastewater inferred by the deconvolution, using the statistical software R [25], and we reported the R^2^. The regressions were performed with data points weighted by the square root of the clinical sample sizes.

## Results

We have developed *LolliPop*, a statistical method to solve the variant deconvolution problem. Our method uses as input the mutation frequencies observed in NGS read data obtained from wastewater samples, and deconvolves into variant relative abundances according to a variant profile matrix and a loss function (Figure 1a). Temporal smoothness is enforced by a fused ridge penalty, enabling deconvolution in the presence of high levels of missing data. The output of Lollipop consists of vectors of relative variant abundance estimates and their confidence intervals, for each time point. These confidence intervals can be computed with a simulation-based approach as well as from a closed-form approximation. Below, we present a comparison of deconvolved relative abundance of variants from the Swiss wastewater surveillance project to clinical data from populations connected to the respective treatment plants. We assess the effect of hyperparameters, and we present confidence bands computed using the different methods introduced. We further assess the robustness of our method on simulated datasets.

**Figure 1:**
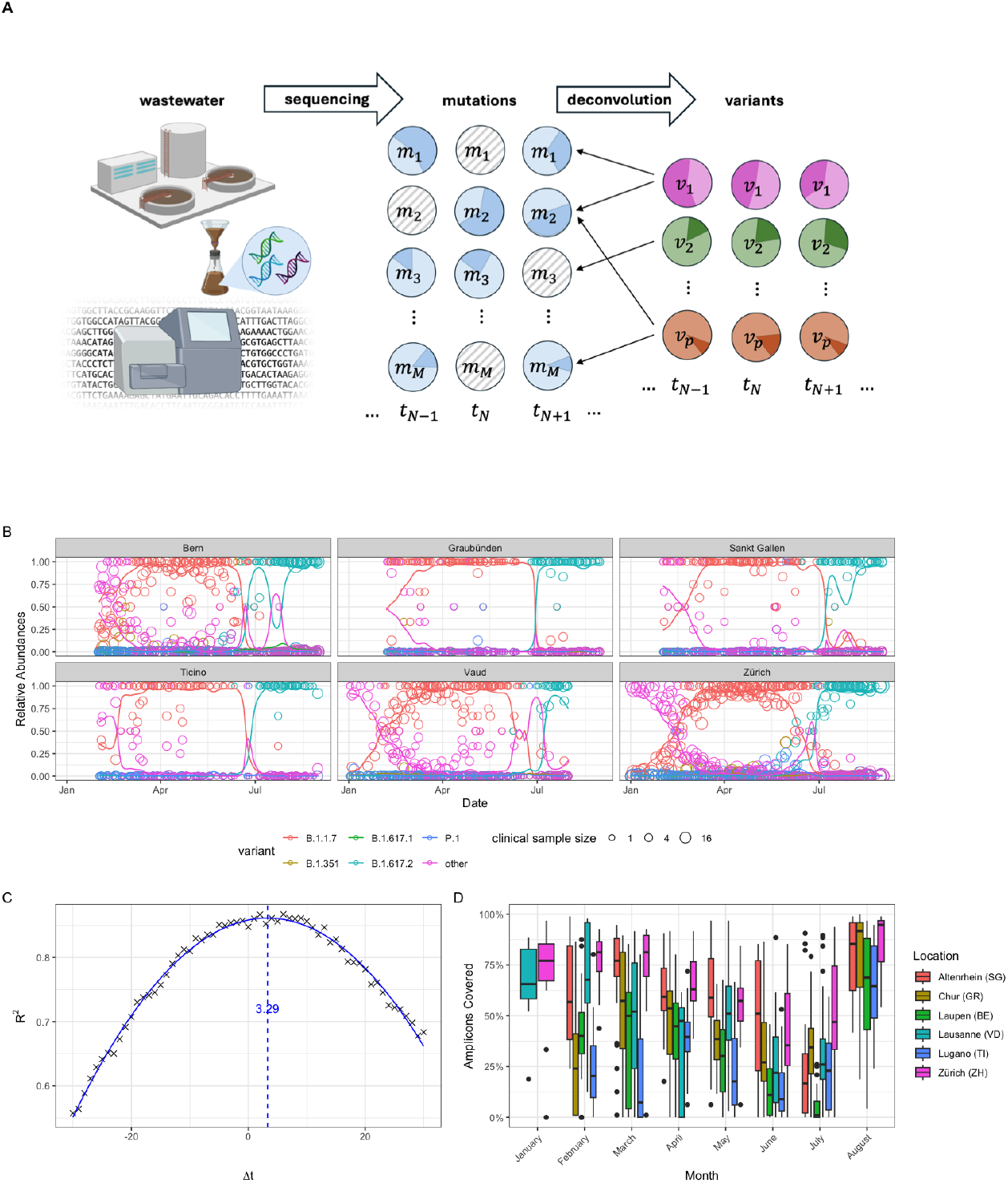
Overview of *Lollipop* for variant deconvolution. **A** Mutation frequencies in wastewater samples of different timepoints are obtained from NGS read counts. The mutation frequencies are deconvolved using the variant definitions, producing estimates of relative variant abundances along with confidence intervals. Repeating the operation for each timepoint tracks the relative abundances of the variants through time. Some values for mutation frequencies are missing (gray), but temporal regularization allows for deconvolution with high levels of missing data. **B** Estimates of variant relative abundances obtained from deconvolution of wastewater data (solid lines), compared to relative abundances of variants in clinical samples of the cantons surrounding the WWTPs (circles). Different colors correspond to the different genomic variants studied. Deconvolution was performed with a Gaussian smoothing kernel with κ = 30 and a scale parameter *α* = 0. 135. **C** Cross-correlation (Pearson *r*) between the deconvolved values of relative abundances and the clinical data estimates weighted by 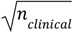 is estimated to peak at 92.8% for Δ*t ≈* 3. 3 days. Crosshairs mark measured cross-correlations, line marks fitted curve. **D** Distribution of the fraction of amplicons covered in the wastewater sequencing data, for each month and for each location. The coverage drops drastically for periods with low incidence (see Supplementary Figure S6).

### Comparison to clinical data

We compared timeseries of relative variant abundances inferred from wastewater sequencing data using *LolliPop* to those estimated using clinical data. To challenge the robustness of *Lollipop*, we used wastewater data including samples from a low incidence period of the pandemic, where viral concentrations were extremely low (Supplementary Figure S6). Despite coverage dropping severely and high dropout rates (Figure 1 d, Supplementary Figure S6), it was still possible to deconvolve the observed mutations into variant relative abundances accurately (Figure 1). We found that the wastewater-based infection dynamics closely follow those derived from clinical sequencing (Figure 1 b). Due to the delay distribution between infection and clinical testing not necessarily matching the delay distribution of shedding and sewage travel time, we expect that the signal in wastewater can be shifted in time compared to clinical samples. The highest cross-correlation value (weighted by root clinical sample size) between wastewater-derived estimates and clinical estimates was *r* = 92.8% estimated for a time lag of Δ*t* = 3. 3 days (Figure 1 c). We performed the deconvolution with kernel bandwidth κ = 30 and a loss scale parameter *α* = 0. 135.

### Confidence intervals

Bootstrapping the mutation counts with subsequent deconvolution was used to produce confidence intervals of the variant relative abundances (Figure 2 a). Alternatively, we also used Wald confidence intervals computed on the logit scale, adjusted for overdispersion and then back transformed (Figure 2 b). Confidence bands from both methods had great overlap (on average, Wald confidence intervals covered ∼89% of the bootstrap confidence intervals), although Wald confidence intervals seemed more conservative. Especially in the Wald confidence intervals, uncertainty was consistently higher during the months in which wastewater samples contained low concentrations of SARS-CoV-2 RNA (June-July) due to low incidence of the virus (Figure 1 d, Supplementary Figure 6).

**Figure 2:**
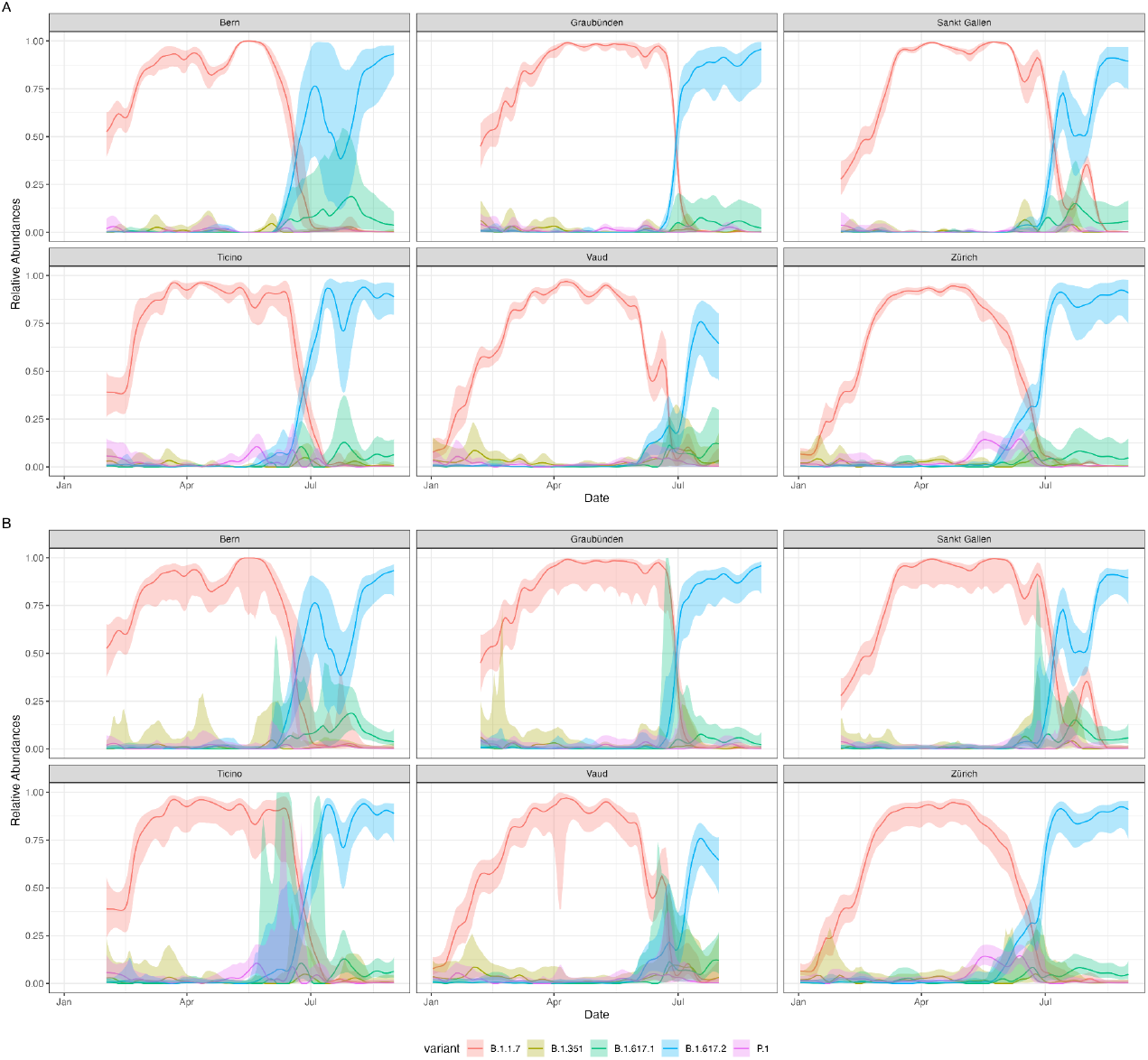
Confidence bands of the deconvolved relative abundance values, at 95% level. **a** Bootstrap confidence intervals computed using 1000 bootstrap samples of the mutations. **b** Wald confidence bands computed using the overdispersion adjusted asymptotic standard error on the logit scale, back-transformed to the linear scale.

### Simulations

The variant deconvolution problem is sensitive to the similarity of variants and the amount of missing values in the sequencing data. To test the limits of the robustness of *Lollipop*, we simulated data from mixtures of variants with varying degrees of similarity and missing data. We found that relative abundances can be estimated robustly, even in the case of very high levels of missing data. This was the case when considering two divergent variants like Alpha and Delta (Figure 3 A, Supplementary Figure S3) as well as when tracking multiple closely related lineages such as Omicron derivatives (Figure 3 b, Supplementary Figure S4). In the absence of simultaneous smoothing, variant deconvolution using the robust SL1 loss displayed higher robustness to missing values as compared to using LS loss. Simultaneous smoothing and deconvolution increased robustness to missing values to a larger extent, at the cost of introducing some bias at the start and end of the time series, a well known artifact of kernel smoothers[26]. The bias introduced by smoothing was generally small and in the presence of missing values by far compensated by the reduced variance of the estimates. Simultaneous smoothing and deconvolution allowed to recover the ground truth time series of variant relative abundances with high accuracy even in the presence of extremely high missing value rates (99%). When considering a mixture of five maximally closely related artificial lineages, simultaneous smoothing offered added accuracy even in the absence of missing values (Figure 3 B, Supplementary Figure S5). Using joint smoothing and deconvolution, it was still possible to recover the time series of relative abundance with high accuracy with missing value rates as high as 88%. Only for the extremely high missing value rate of 99%, deconvolution started to fail and only very rough estimates of the average relative abundances of the variants could be recovered.

**Figure 3:**
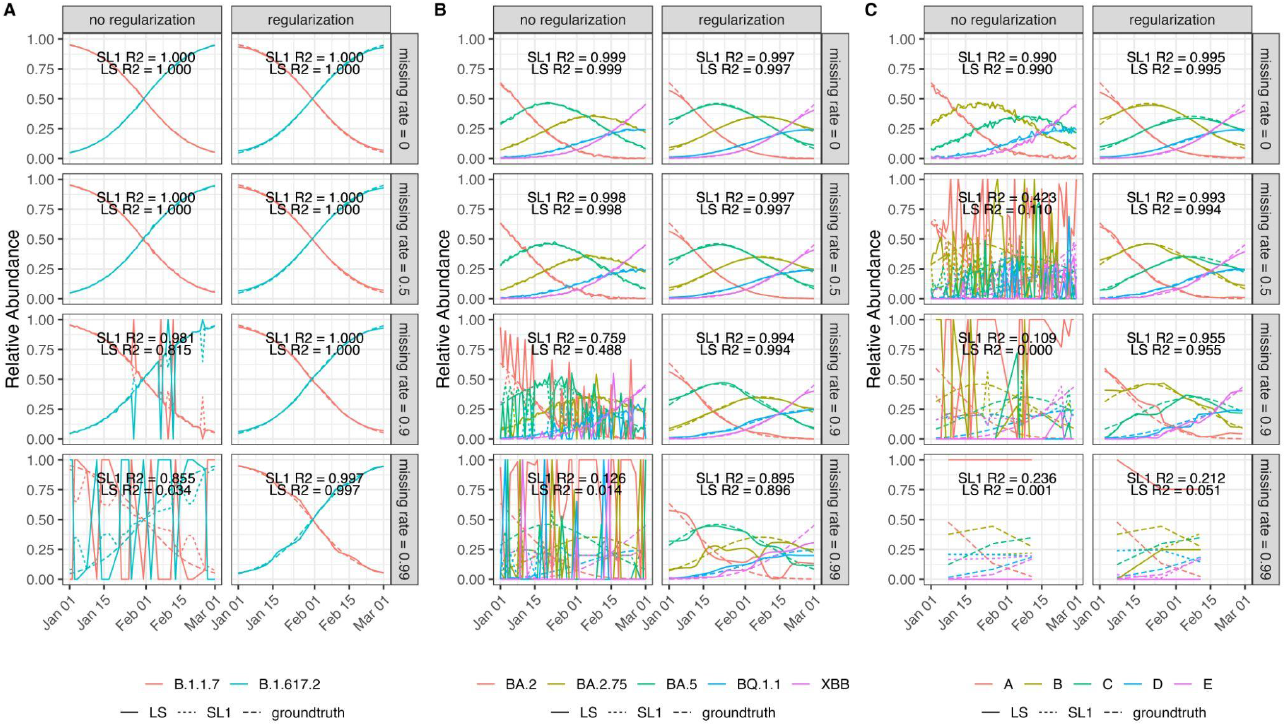
Simulation experiments displaying the robustness conferred by regularization to varying levels of missing data (0%, 50%, 90% and 99%) on a 60-timestep timeseries of variant competition for different combinations of variants. Dashed lines represent simulated ground truth. Solid lines and dotted lines show deconvolution results with the LS (i.e., SL1 with *α* = 1) and SL1 (*α* = 0. 13) loss functions, respectively, with annotations showing their respective R^2^ values compared to ground truth. **a** Simulated time series of B.1.617.2 taking over B.1.1.7. **b** Simulated time series of highly similar Omicron subvariants. **c** Simulated time series of a mixture of five artificial variants designed to be maximally similar.

### Hyperparameters

We ran *LolliPop* on simulated data consisting of a mixture of five closely related Omicron subvariants, using a grid of hyperparameter values. These tests were performed under varying levels of missing data. Increasing the smoothing bandwidth κ from zero initially reduced the RMSE, but it gradually increased again at higher values due to bias introduced by oversmoothing (Supplementary Figure S1). The benefit of using a nonzero smoothing bandwidth was greater in the presence of high levels of missing data. In contrast, the choice of the loss scale parameter *α* (which controls the breakpoint between *l*_2_ and *l*_1_ loss) had a negligible effect on RMSE.

Next, we applied LolliPop to the Swiss wastewater monitoring data from the two largest WWTPs, again using a grid of hyperparameter values. Increasing the smoothing bandwidth κ improved the goodness of fit with clinical sequencing data, as measured by R^2^ (Supplementary Figure S2). For the loss scale parameter *α*, the best fits were obtained for values between 0.1 and 0.25. Based on these results, we recommend using a smoothing bandwidth greater than 10 days and a loss scale parameter between 0.1 and 0.25.

### Runtime

Wall time on a MacBook Air M1 2020 was ∼2s for deconvolving all 1295 wastewater samples (225 timepoints, 6 locations, 5 variants defined through 94 mutations) using the LS loss function (i.e., SL1 with *α* = 1). Using the SL1 loss function, wall time was ∼1min for the same task. Wall time was ∼1min for reparameterized Wald confidence intervals. Constructing confidence intervals from 1000 bootstrap samples) had a wall time of ∼30min. All computations were performed using a single CPU core.

## Discussion

We have presented *LolliPop*, a method for solving the variant deconvolution problem. Deconvolving patterns of mutations in wastewater NGS data is necessary to track the relative abundance of viral variants. We showed its application to data from the Swiss wastewater monitoring program, extending over eight months at six different locations around the country. We found that the deconvolved values of relative abundance closely follow the dynamics observed in the clinical sequencing effort, even in the presence of high rates of missing values. We further evaluated the robustness to missing values of our method on simulated time series of increasing complexity. We found that ground truth values can be recovered with high accuracy in the presence of high and very high rates of missing values.

Regarding runtime, we observed that *LolliPop* can be easily used on a standard laptop computer to process thousands of samples in a matter of seconds or minutes, using a single computing core. This emphasizes how the program can readily be used in analyses that scale up to national monitoring programs.

The main limitation of *LolliPop* is that it relies on compiling variant definitions from repositories of clinical samples. It therefore does not allow for epidemiological surveillance based on wastewater alone, without any supplementing clinical sequencing. In practice, only one positive sample of a given variant is needed to provide a full haplotype, which can then be used as the profile of a variant to track its relative abundance in wastewater. *Lollipop* is therefore most effective in an integrated genomic surveillance program. Another limitation is that the current implementation considers genomic positions independently, which can lower the sensitivity of the detection for very low abundance variants. *Lollipop* could in principle accept local haplotype counts (e.g., derived from read-pairs profiles) instead of mutation counts, which could alleviate this issue.

Modifications to *LolliPop* can be easily implemented. First, the behavior of the deconvolution under different types of loss functions could be investigated. We have used here the Soft *l*_1_ loss, and the results were not very strongly affected by the choice of the scale parameter *α*, indicating robustness of the method to this choice. Other types of losses could certainly have interesting properties for the problem at hand. For example, if the number of candidate lineages to deconvolve grows, building in a sparsity assumption by adding an *l*_1_ regularization term of the relative variant abundances could lower the variance of their estimates.

Another component that can readily be modified is the type of kernel used for enforcing temporal regularization. We have used a Gaussian kernel, but other choices could be relevant depending on the application. For example, kernels elicited on expert knowledge could be used for more precisely relating the relative abundances in samples to the relative incidences in the population. The temporal dynamics of viral shedding in wastewater are generally described by a shedding load distribution [27], which could be accounted for by an asymmetric and non zero-centered kernel. In general, we recommend using a kernel with sufficient bandwidth to robustify the method against missing values.

To assess the uncertainty in the estimates of relative abundances, we have derived a bootstrap-based and an analytical approach to produce confidence intervals. Bootstrap confidence intervals are easier to derive but are by design computationally intensive, whereas analytical expressions for confidence intervals can provide substantial speedup. The Wald confidence intervals we derived here include three components of the variance of the estimates: one to account for the mutation overlap between variant definitions, one to account for the quadratic form of the variance of a binomial sampling, and one to account for overdispersion in read counts. The Wald confidence intervals computed on the linear scale suffers from known shortcomings, as it can exit the [0,1] range, which is why we compute them on the logit scale. In our results, the Wald confidence intervals very closely resemble the bootstrap confidence intervals, while providing an almost 30x speedup in computation time.

*Lollipop* has built in the assumption of temporal continuity of the variant relative abundances in the form of kernel smoothing simultaneously with deconvolution, which enforces a ridge penalty on the temporal variation of the relative abundance. As in a monitoring program, usually multiple locations are being monitored, another useful assumption to build in the deconvolution could be that of spatial continuity if the spatial resolution allows for it. Jointly smoothing the different locations might offer increased robustness to the estimates by partial pooling of the information.

To summarize, *LolliPop* solves the variant deconvolution problem, taking into account the time series nature of wastewater sequencing datasets and mitigates the extremely high levels of noise and dropouts these experiments typically display. Our method can estimate uncertainty using different approaches, including analytical confidence bands with short computation times. As such, *LolliPop* enables genomic variant tracking in large-scale wastewater-based epidemiology projects.

## Data Availability

All data and code used to produce the results presented in this article is available at https://doi.org/10.5281/zenodo.15277339.

## Acknowledgements

This study was supported by the Swiss National Science Foundation (grant no. CRSII5_205933). We thank all members of the Wastewater-based Infectious disease Surveillance (WISE) consortium.

## Author contributions

Conceptualization: D.D, N.B. Methodology: D.D, I.T, P.I.B. Software: D.D, I.T, P.I.B. Validation: D.D. Formal analysis: D.D. Investigation: D.D. Resources: N.B. Data curation: D.D, I.T. Writing – original draft: D.D. Writing – review and editing: all authors. Visualization: D.D. Funding acquisition: D.D, N.B. Supervision: N.B.

## Footnotes

1 https://bsse.ethz.ch/cbg/research/computational-virology/sarscov2-variants-wastewater-surveillance.html

