## Supplementary Materials for "Tracking SARS-CoV-2 genomic variants in wastewater sequencing data with *LolliPop*"

### Supplementary Material for “Tracking SARS-CoV-2 genomic variants in wastewater sequencing data with *LolliPop*”

#### Supplementary 1

Let $b$ be the vector of variant relative abundances, $\pi$ be the vector of expected fractions of mutated reads given a design matrix $X$, i.e., $\pi(b) = Xb$. The Hessian matrix of the deconvolution likelihood model is found as follows. We differentiate twice the log-likelihood of the model

$$\frac{\partial}{\partial b}\ell(\pi) = \frac{\partial}{\partial \pi(b)}\ell(\pi(b))\frac{\partial}{\partial b}\pi(b)$$

$$\Rightarrow \frac{\partial^2}{\partial b^2}\ell(\pi(b)) = \frac{\partial}{\partial b}\left\{\frac{\partial}{\partial \pi(b)}\ell(\pi(b))\right\}\frac{\partial}{\partial b}\pi(b) + \underbrace{\frac{\partial^2}{\partial b^2}\pi(b)\frac{\partial}{\partial \pi(b)}\ell(\pi(b))}_{\pi(b)=Xb\Rightarrow\frac{\partial^2}{\partial b^2}\pi(b)=0}$$

$$= \frac{\partial}{\partial b}\frac{\partial}{\partial \pi(b)}\ell(\pi(b))\frac{\partial}{\partial b}\pi(b) \quad = \left(\frac{\partial}{\partial \pi(b)}\frac{\partial}{\partial b}\ell(\pi(b))\right)^{\top}\frac{\partial}{\partial b}\pi(b)$$

$$= \left(\frac{\partial}{\partial \pi(b)}\frac{\partial}{\partial \pi(b)}\ell(\pi(b))\frac{\partial}{\partial b}\pi(b)\right)^{\top}\frac{\partial}{\partial b}\pi(b) \quad = \left(\frac{\partial}{\partial b}\pi(b)\right)^{\top}\underbrace{\left(\frac{\partial^2}{\partial \pi(b)^2}\ell(\pi(b))\right)^{\top}}_{=\frac{\partial^2}{\partial \pi(b)^2}\ell(\pi(b))}\frac{\partial}{\partial b}\pi(b)$$

The Fisher information matrix is then

$$I_b(b) = E\left[-\frac{\partial^2}{\partial b^2}\ell(\pi)\right] = E\left[\left(\frac{\partial}{\partial b}\pi\right)^{\top}\left(-\frac{\partial^2}{\partial \pi^2}\ell(\pi)\right)^{\top}\frac{\partial}{\partial b}\pi\right] = X^{\top}diag\left\{\pi_m(1-\pi)_m\right\}^{-1}_{m=1,\dots,M}X$$

#### Supplementary Figures

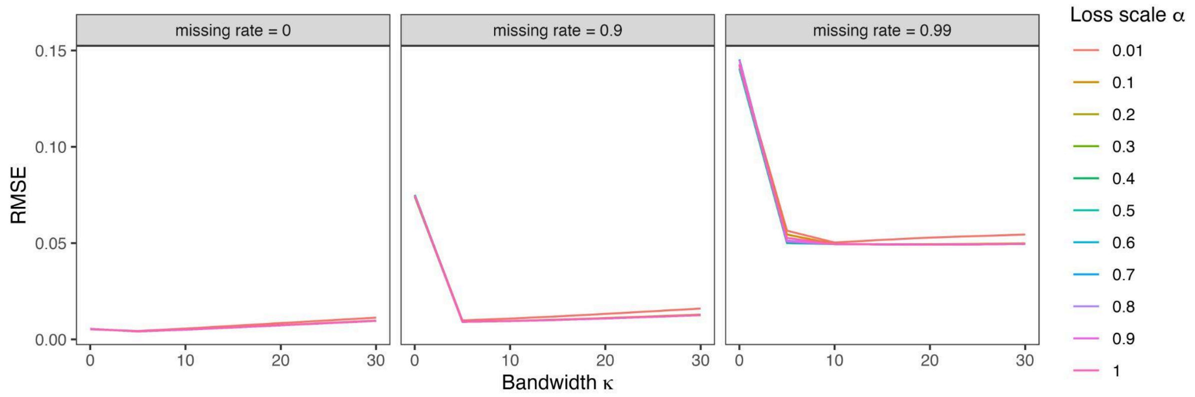


**Figure S1:** Root Mean Square Error (RMSE) of the deconvolved simulated data, as a function of the kernel bandwidth κ and α scale parameters of the robust regression. The accuracy of the deconvolution was measured on the simulated data shown in Supplementary Figure S4.

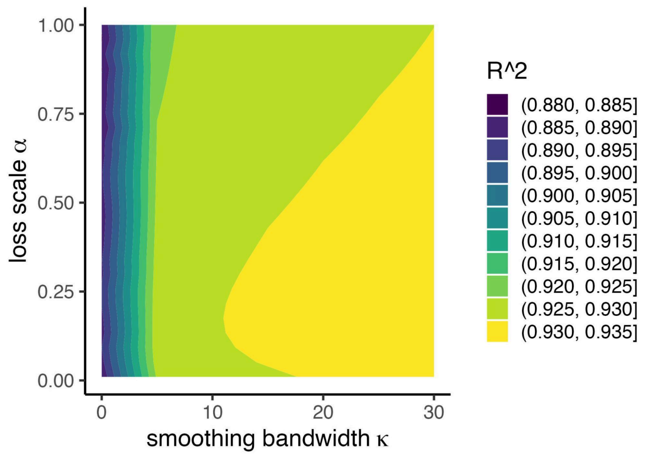


**Figure S2:** Goodness of fit of the robust kernel deconvolution of the wastewater NGS, as a function of the kernel bandwidth $\kappa$ and $\alpha$ scale parameters of the robust regression. Goodness of fit is evaluated by regressing on estimates of relative abundances of variants obtained from clinical sequencing data.

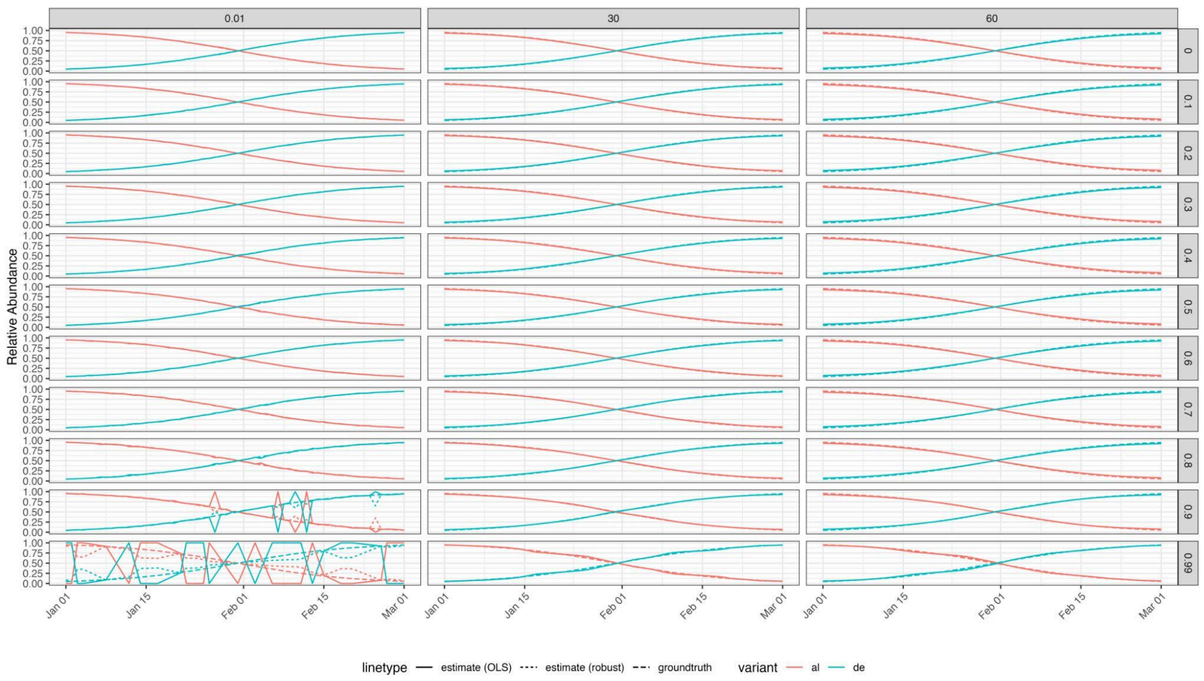


**Figure S3:** Simulation experiments, displaying the effect of kernel smoothing on varying levels of missing value on a 60-timesteps timeseries of B.1.617.2 (de) taking over B.1.1.7. (al). Columns are different values of the smoothing bandwidth, rows are different levels of missing values. Dashed lines represent ground truth, solid lines and dotted lines show deconvolution results with the LS and SL1 loss functions, respectively.

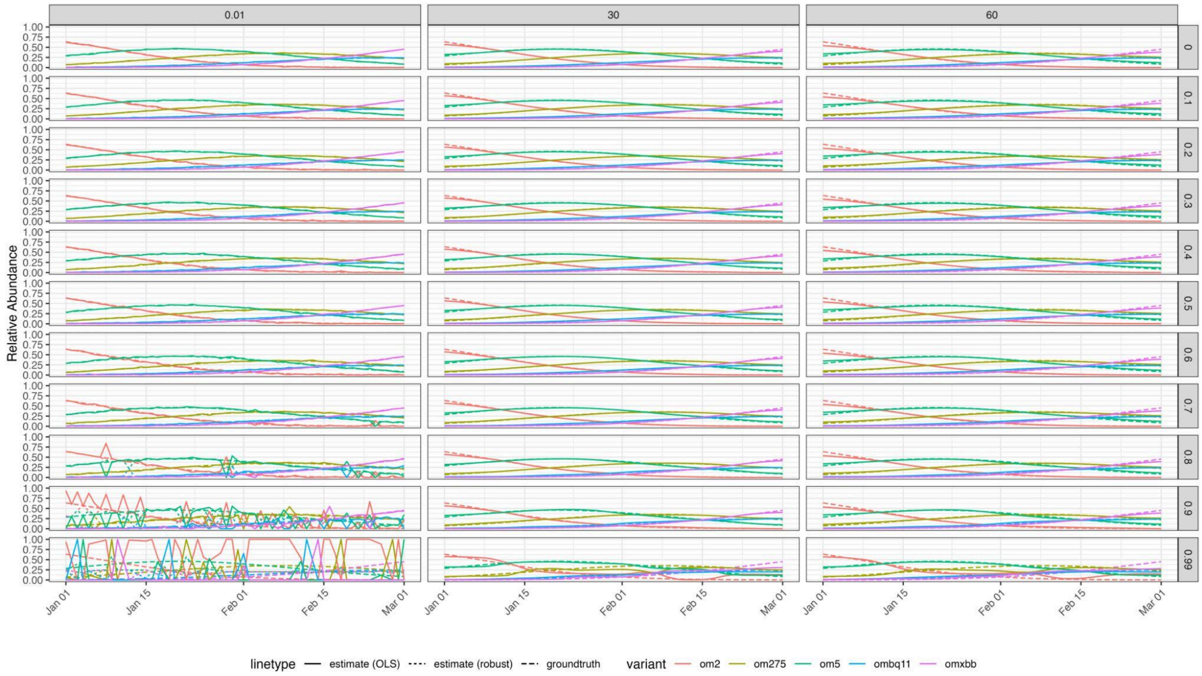


**Figure S4:** Simulation experiments, displaying the effect of kernel smoothing on varying levels of missing value on a 60 timesteps time series of the five closely related Omicron derivatives BA.2 (om2), BA.2.75 (om275), BA.5 (om5), BQ.1.1 (ombq11) and the recombinant XBB (omxbb). Columns are different values of the smoothing bandwidth, rows are different levels of missing values. Dashed lines represent ground truth, solid lines and dotted lines show deconvolution results with the LS and SL1 loss functions, respectively.

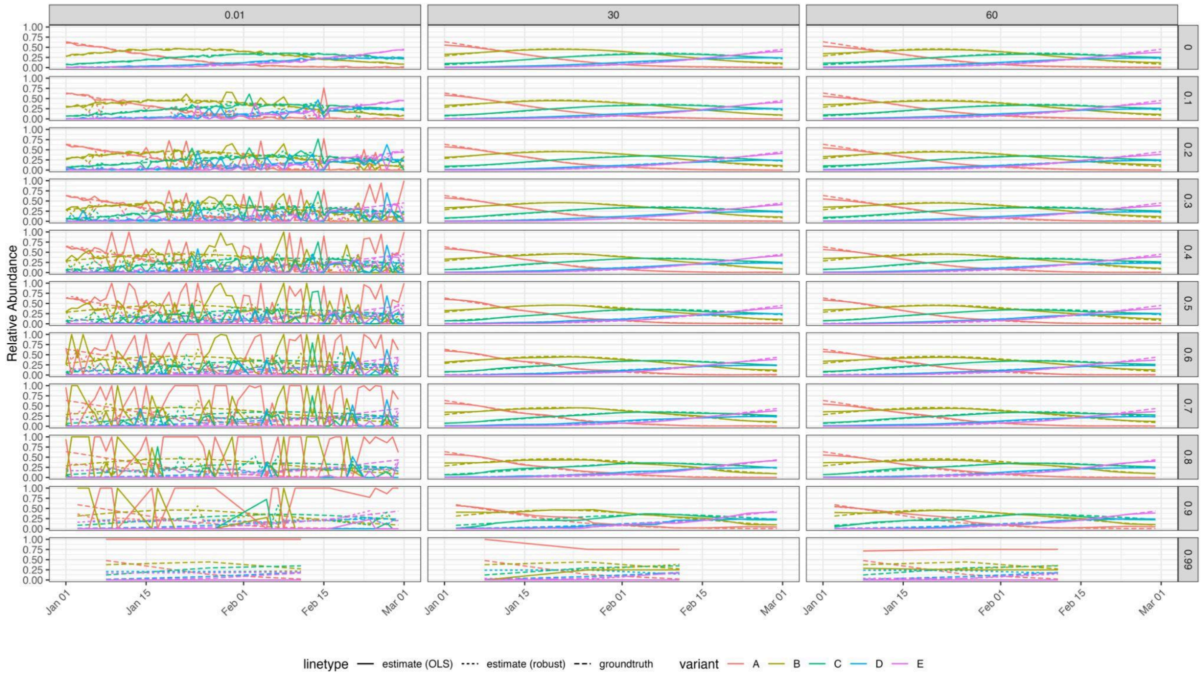


**Figure S5:** Simulation experiments, displaying the effect of kernel smoothing on varying levels of missing value on a 60 timesteps time series of the five artificially closely related variants. Columns are different values of the smoothing bandwidth, rows are different levels of missing values. Dashed lines represent ground truth, solid lines and dotted lines show deconvolution results with the LS and SL1 loss functions, respectively.

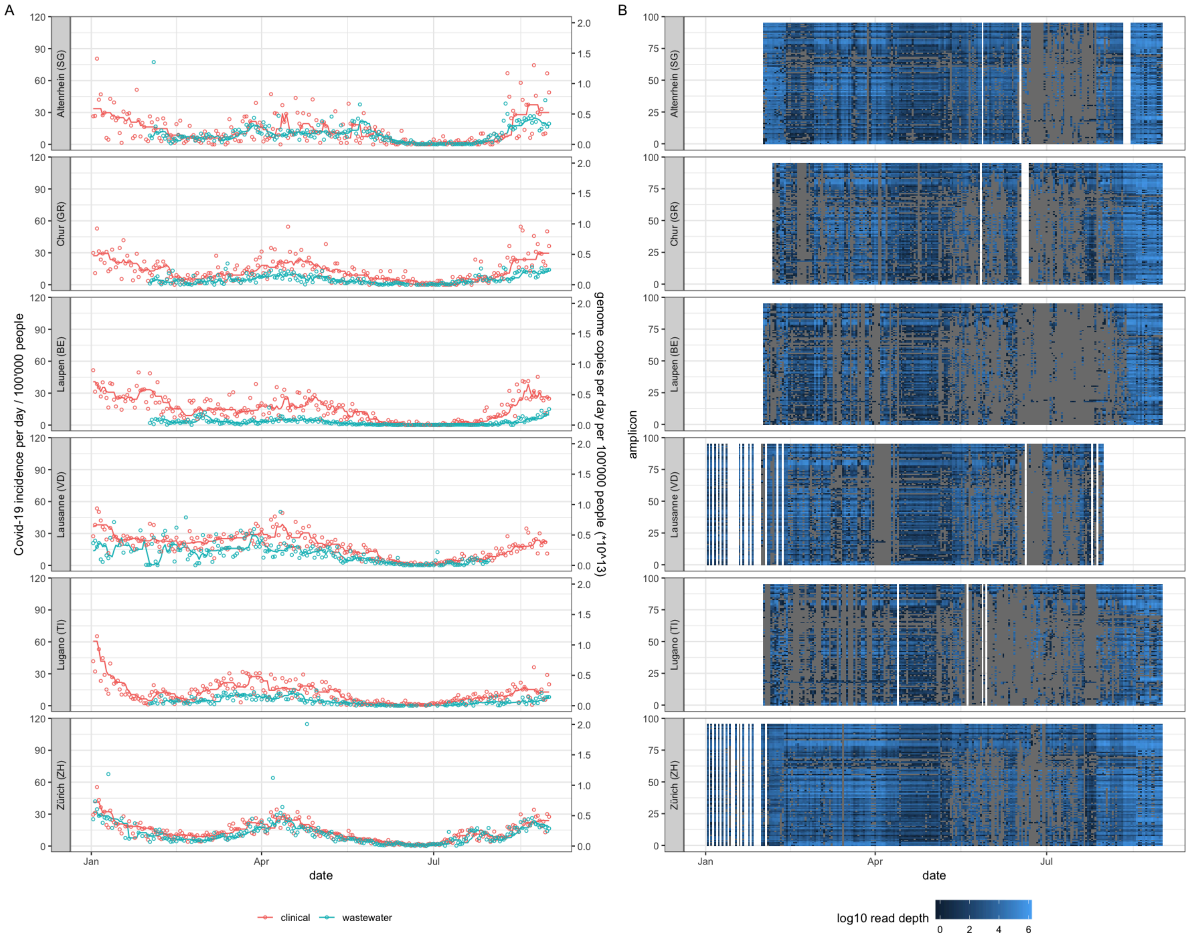


**Figure S6:** Viral loads in the wastewater samples and daily incidence in the catchment area of the treatment plants. Points are measured values and lines are 7-day median values. Data from https://sensors-eawag.ch/sars/overview.html (A). $Log_{10}$ read depth per amplicon, per sample across the different locations. Gray values represent amplicons with zero coverage, i.e. dropouts (B).
